# Natural History of Prenatally Identified Children with 48,XXYY Syndrome in Infancy and Early Childhood

**DOI:** 10.64898/2026.06.04.26353909

**Authors:** Kayla Nocon, Karli Swenson, Samantha Bothwell, Susan Howell, Shanlee Davis, Chijioke Ikomi, Judith Ross, Nicole Tartaglia

**Affiliations:** Department of Pediatrics, University of Colorado School of Medicine, Aurora, Colorado, USA; eXtraordinarY Kids Program, Children’s Hospital Colorado, Aurora, Colorado, USA; Nemours Children’s Hospital-Delaware, Wilmington, DE, USA

**Author notes:** Denotes co-first authors (Kayla Nocon, Karli Swenson). **Address correspondence to**: Nicole Tartaglia, MD, Developmental Pediatrics, 13123 East 16^th^ Ave, B140. **Role of Funder/Sponsor:** The NIH had no role in the design and conduct of the study. Contents are the authors’ sole responsibility and do not necessarily represent official NIH views. **Clinical Trial Registration and Data Sharing Statement (if any)**: This study is registered on ClinicalTrials.gov NCT03396562 https://clinicaltrials.gov/study/NCT03396562. Data sharing is available through requests from NICHD DASH (https://dash.nichd.nih.gov/) and includes deidentified individual participant data, study protocols, DASH data codebook, and the informed consent form. The data will be made available to researchers who provide methodologically sound proposals approved by the NICHD DASH Data or Biospecimen Access Committee. **Article Summary:** We present medical features of a cohort of children with 48,XXYYidentified via noninvasive prenatal screening, including congenital, medical, and developmental features in comparisons to male trisomies 47,XXY and 47,XYY. **What’s Known on This Subject:** 48,XXYY occurs in approximately 1 in 18,000 – 1 in 40,000 male births and has been traditionally diagnosed in childhood or adolescence secondary to congenital anomalies, developmental delays, autism, or hypogonadism. Multisystem involvement necessitates neurodevelopmental and medical intervention and support. **What This Study Adds:** This prospective observational study characterizes medical and developmental features of infants with prenatally identified 48,XXYY from infancy through the first years of life. We compare these findings to historical postnatally identified cohorts, providing more accurate data for prenatal counseling and childhood surveillance.

## Abstract

**Background:** 48,XXYY syndrome is a rare sex chromosome aneuploidy (SCA) characterized by neurodevelopmental deficits and medical comorbidities. The limited information available in the literature is almost exclusively limited to postnatally diagnosed cases. This study aims to describe the early medical and developmental features of prenatally identified 48,XXYY infants, with comparisons to 47,XYY, 47,XXY cohorts, and typical populations, as well as previously reported postnatally diagnosed 48,XXYY cases.

**Methods:** The eXtraordinarY Babies Study prospectively follows children prenatally identified to be at high risk for SCA with annual medical and neurodevelopmental evaluations. Data presented herein include the prevalence of medical conditions, developmental milestones, developmental and adaptive functioning assessment scores, and therapy utilization in participants confirmed to have 48,XXYY. Comparisons were made between this cohort and the typical population, infants with 47,XYY and 47,XXY also enrolled in the eXtraordinarY Babies Study, and a 2008 cohort of individuals postnatally identified 48,XXYY.

**Results:** Infants with 48,XXYY exhibited a range of early medical features, including high rates of feeding and GI disorders (breastfeeding difficulties, gastroesophageal reflux, and eosinophilic esophagitis), allergic disorders (food allergies and environmental allergies), and hypotonia. Developmental and adaptive functioning scores indicated delays in motor, communication, and social domains, with nearly all infants receiving speech therapy, physical and/or occupational therapy. Comparisons with the 47,XYY and 47,XXY cohorts revealed more medical and developmental challenges in the 48,XXYY group, however there was variability and some overlap with both the general population and sex chromosome trisomy conditions. Additionally, comparison to the 2008 postnatally identified 48,XXYY cohort indicated that while prenatal diagnosis allowed for earlier intervention, developmental outcomes in the first years of life were similar between the two groups.

**Conclusions:** 48,XXYY diagnosed prenatally facilitates early monitoring, anticipatory guidance, and proactive referrals for medical evaluations and intervention, given developmental delays and medical challenges are more common in infancy and early childhood compared to the general population and trisomy SCAs. These findings provide valuable insights for genetic counselors and healthcare providers, emphasizing the spectrum of medical and developmental findings and importance of early and proactive care to support individual outcomes. Prospective study of this prenatally identified cohort will provide important natural history and phenotypic variability in XXYY, as well as identification of predictors of health and developmental outcomes.

## Introduction

48,XXYY syndrome is a rare sex chromosome aneuploidy (SCA) that occurs in approximately 1 in 18,000 to 1 in 40,000 male births.^1^ This condition arises from nondisjunction errors resulting in the presence of an extra X and Y chromosome, leading to a 48,XXYY karyotype characterized by clinical features that impact neurodevelopment, endocrine function, and physical health.^2^ Historically, much of the literature on 48,XXYY has focused on individuals diagnosed postnatally secondary to congenital malformations, developmental delays, autism, features associated with hypogonadism (small testes, testosterone deficiency), or medical problems such as seizures that prompted genetic testing.^2-4^ As a result, very little prospective data are available for the very early years, or for individuals with incidental diagnoses.

Previous literature on 48,XXYY syndrome has documented increased risk for a range of medical and neurodevelopmental diagnoses. Common findings include developmental delays, cognitive / learning impairments, speech disorders, and motor skill deficits.^2,4,5^ Neurodevelopmental disorders such as autism spectrum disorder (ASD) and attention-deficit/hyperactivity disorder (ADHD) are also frequently reported, with a significant proportion of individuals displaying behavioral challenges, including impulsivity, anxiety, and social skills difficulties.^2,6^ XXYY syndrome is often classified as a variant of Klinefelter syndrome (47,XXY) due to the shared features of tall stature, hypogonadism, and increased risk for diagnoses such as ASD. However, as there is additional gene dysregulation and overexpression due to having four sex chromosomes instead of the three present in Klinefelter syndrome, we herein analyze those with 48,XXYY as a distinct population.

Many males with 48,XXYY have tall stature, along with increased risk for non-specific physical features such as facial dysmorphisms and skeletal anomalies such as clinodactyly, pes planus, radioulnar synostosis, or joint hypermobility.^2,7^ Endocrinological abnormalities are nearly universal in men with 48,XXYY, with testicular dysfunction leading to microcorchidism, hypergonadotropic hypogonadism (presenting as incomplete puberty, low testosterone levels, gynecomastia), and infertility.^8,9^ In addition, congenital and acquired cardiovascular and metabolic issues are common, with elevated risks of congenital heart defects, venous thromboembolism, and type 2 diabetes.^2,10^ Immunological problems, including recurrent respiratory infections, asthma, and eosinophilic esophagitis have also been noted, as well as neurological conditions such as epilepsy and tremor.^2,11^

Previous studies, largely based on individuals with a postnatal diagnosis, highlight the importance of early detection and multidisciplinary management to address the complex health needs of individuals with 48,XXYY syndrome. As prenatal cell-free DNA (cfDNA) screening becomes widely adopted as standard of obstetric care, a growing population of infants ascertained with SCAs, including 48,XXYY, has followed.^12^ Subsequently, historic ascertainment bias attributed to a more severe phenotypic presentation will be challenged, as findings from a prenatally identified cohort of infants provides a less-biased sample for comparison.^13^ This allows researchers to include and report the full spectrum of medical problems associated with 48,XXYY, including milder cases that might have gone undiagnosed previously, thereby offering a clearer understanding of the condition’s natural history and variability in medical outcomes.

This case series describes 15 males ranging from 12-months to 8-years old prenatally identified to have a high risk for SCA and postnatally confirmed 48,XXYY syndrome who have been followed prospectively since their first year of life. We describe the most common physical findings, medical conditions, and developmental trajectories across the first years of life using standardized parental questionnaires and direct assessments. We further compare these findings to prospectively followed 47,XXY and 47,XYY cohorts and previous reports of 48,XXYY to allow contextualization of results and application for counseling and management.

## Methods

### Study Design, Recruitment, and Ethical Compliance

Data for this report were collected as part of a longitudinal natural history study of early medical and developmental features in children with prenatally diagnosed sex chromosome trisomy (SCT) called the eXtraordinarY Babies Study.^14^ Participants were recruited through national advocacy organizations (The XXYY Project, Association for X and Y Chromosome Variations), prenatal genetic testing companies, genetic counselors, professional conferences, and social media websites. Data were collected between September 2017 and January 2026. Inclusion criteria were prenatal identification of SCA, confirmatory karyotype, enrollment prior to 12 months of age, and English or Spanish speaking. Participants were excluded if they had an additional diagnosis of a different genetic or metabolic disorder with neurodevelopmental or endocrine involvement, prematurity of <34 weeks, and/or a complex congenital malformation not previously associated with SCT. This study was approved by the Colorado Multiple Institutional Review Board (COMIRB #17-0118), and all parents provided written consent for themselves and their children per ethical requirements. This project is funded by NICHD (ClinicalTrials.gov NCT03396562) and is being conducted at two sites including University of Colorado / Children’s Hospital Colorado and Nemours Children’s Hospital Delaware.

### Data Collection

#### Procedures

Study visits occur at 2-months, 6-months, and 12-months of age and continue annually until the age of 10, with in person assessments occurring at 12-months, 3-years, 6-years, 8-years, and 10-years of age and telehealth visits at other timepoints. All visits include a battery of questionnaires completed prior to the visits, which are then validated through interview with the study physicians. In-person visits also include an extensive battery of neurodevelopmental testing, physical examination, and biosample collection. Electronic medical records are reviewed to gather additional data. Data were electronically stored and managed using REDCap, hosted by the University of Colorado.

#### Perinatal, Medical and Family History

Following enrollment, parents completed questionnaires assessing prenatal and birth history, including pregnancy details, diagnostic history, and birth information. Parents also completed a comprehensive health and developmental history questionnaire covering medical diagnoses, medications, hospitalizations, surgeries, imaging and diagnostic testing, developmental diagnoses, milestone attainment, therapies, educational supports, and family health updates. Additional questionnaires collected detailed information on family history, including medical, learning, neurodevelopmental, and mental health conditions in family members, as well as demographic characteristics including race, ethnicity, parental education, occupation, and income. During study visits, clinicians conduct an interview with the parent(s) to review and verify survey responses.

#### Developmental History

Parents were asked to report if their child had achieved key developmental milestones, including eight gross motor skills and four expressive communication milestones. These milestones were chosen as they are easily observed by parents in a natural setting and have normative data available. Response options were “Yes”, “No”, “I don’t know”. Parents who marked “Yes” were prompted to estimate the age in months the child achieved this skill. Responses were verified by study clinicians through interviews with parents. The twelve developmental milestones collected for the study were compared with existing published norms. Milestone percentiles from normative data were included from the Denver II Scales, the World Health Organization (WHO) Motor Development Study, and the Primitive Reflex Profile (PRP).^15-17^

#### Vineland-3

Vineland-3 is an instrument completed by the child’s parent/primary caregiver to assess adaptive skills across several domains (Communication, Daily Living, Socialization, Motor), resulting in an Adaptive Behavior Composite score.^18^ This instrument can be done as an in-person interview with a clinician during the time of the study visit or completed electronically by the parent/primary caregiver prior to the study visit. Vineland-3 subtests are validated with mean V-scores of 15 and a standard deviation of 3 and the adaptive behavior composite score has a mean of 100 with a standard deviation of 10. Participants were administered the Vineland-3 for in-person and telehealth visits (ages 2, 6, 12, 24, 36, 48-months, and 6, 8 and 10 years old). After the instrument was administered, scores were calculated in Q-global and stored in REDCap.

#### Bayley Scales of Infant and Toddler Development, 3rd Edition (Bayley-III)

The Bayley-III was used to directly assess child development across cognitive, language (expressive and receptive), and motor (fine and gross) domains. The Bayley-III is commonly used for assessment of infants and toddlers (age 1-42 months) in both clinic and research settings and has excellent reliability and validity.^19,20^ Bayley-III subtests are normed by age with mean scaled scores of 10 and a standard deviation of 3. Participants were administered the Bayley-III at each in-person visit (ages 2, 6, 12 and 36-months).

### Statistical Analysis

Medical and developmental features of the 48,XXYY cohort were compared to the general population, previous literature on postnatally identified 48,XXYY individuals, as well as prenatally identified 47,XXY and 47,XYY participants of the eXtraordinarY Babies Study. Due to the small sample size, Fisher’s Exact tests were utilized to determine if the prenatally identified 48,XXYY had higher prevalences of outcomes than the comparison groups. The distribution of current age was not significantly different between the 48,XXYY cohort and the 47,XXY and 47,XYY cohorts so age was not controlled for in analyses. All prevalences are summarized as N(%).

Developmental milestone ages, Vineland-3 V-scores, and Bayley-III scaled scores for 48,XXYY participants were summarized as Median [Interquartile Range], due to non-normality as assessed with Shapiro Wilk tests, and compared to 47,XXY and 47,XYY participants using Wilcoxon-Rank sum tests. The distribution of milestone ages is compared to normative data, taken from the Denver II scales, the World Health Organization (WHO) Motor Development Study, and the Primitive Reflex Profile (PRP).^15-17^ As normative data were only presented as percentiles, we simulate a normative distribution based on the provided percentiles for comparison under the assumption of a non-normal distribution. Continuous demographics are either presented as mean ± standard deviation or median [interquartile range], depending on normality assessed by Shapiro-Wilk tests. Analyses were performed in R, version 4.4.0. As this is predominantly a small sample case series analysis, an unadjusted significance threshold of 0.05 was applied, with a False Discovery Rate (FDR) p-value adjustment made for comparison of medical finding prevalences across cohorts. Missing data were verified as true missingness and complete case analysis was performed under the assumption of missing at random.

## Results

As of January 2026, the median age of participants enrolled in the eXtraordinarY babies study with 48,XXYY was 61 months (∼5 years) and had been in the study for a median of 4.3 years (Table 1). Most participants (14; 93.3%) had prenatal cfDNA results suggesting an increased risk for a sex chromosome trisomy (47,XXY or 47,XYY). Most families (80%) declined prenatal diagnostic confirmation (amniocentesis) and were diagnosed with 48,XXYY after birth via karyotype (60%), microarray (6.7%), or a multiple genetic tests (26.7%).

**Table 1.** Prenatally Identified 48,XXYY Cohort Demographics (N=15)

|  |  |
| --- | --- |
| **Age at Enrollment (Months)** | 6.3 ± 4.5 |
| **Age at Last Visit (Months)** | 67.6 ± 25.2 |
| **Duration of follow up (Months)** | 51.6 ± 25.1 |
| **Race** |  |
| White | 13 (86.7%) |
| African American or Black | 1 (6.7%) |
| Multiple Race | 1 (6.7%) |
| **Ethnicity** |  |
| Hispanic/Latino | 1 (7%) |
| Not Hispanic/Latino | 14 (93.3%) |
| **Annual Household Income** |  |
| < \$75,000 | 3 (20%) |
| \$75,000-150,000 | 5 (33.3%) |
| \$150,000-250,000 | 5 (33.3%) |
| >\$250,000 | 2 (13.3%) |
| **Highest parent education<sup>1</sup>** |  |
| High School Graduate | 1 (6.7%) |
| Partial College | 1 (6.7%) |
| Standard College/University Degree | 2 (13.3%) |
| Graduate/Professional Degree | 11 (73.3%) |
| **Birth History** |  |
| Full Term ($\geq 37$ weeks) | 14 (93.3%) |
| Gestational Age (weeks) Mean ± SD | 38.4 ± 1.1 |
| Birth Weight (kg) Mean ± SD | 3.0 ± 0.5 |
| Maternal Age (years) Mean ± SD | 32.6 ± 5.5 |
| **Prenatal cfDNA Results** |  |
| cfDNA + 47,XXY | 12 (80%) |
| cfDNA + 47,XYY | 2 (13.3%) |
| cfDNA – Indeterminate | 1 (6.7%) |
| **Indication for prenatal screening / testing<sup>2</sup>** |  |
| Routine/Elective screening | 12 (80.0%) |
| Advanced Maternal Age | 2 (13.3%) |
| Abnormal Ultrasound | 1 (6.7%) |
| **Confirmatory Genetic Testing<sup>3</sup>** |  |
| Amniocentesis only | 1 (6.7%) |
| Amniocentesis + Postnatal Testing | 2 (13.3%) |
| Postnatal Testing Only | 12 (80%) |
| **Physical / Medical Features** |  |
| 12 Month Length Z-score | -0.82 [-1.51, -0.07] |
| 12 Month Weight Z-score | -0.25 [-0.61, 0.13] |
| Previous testosterone treatment | 4 (26.7%) |

Data are presented as median and [IQR] or N (%)
<sup>1</sup> Parental education is taken as the maximum education level reported between both parents. If only one parental education level was reported, that education level was used.
<sup>2</sup> Abnormal Ultrasound identified possible heart defect and corpus callosum defect.
<sup>3</sup> 2 of the 3 participants who had an amniocentesis following a positive cfDNA also had postnatal confirmation, while one with positive cfDNA and amniocentesis did not have postnatal testing.
Summaries represent N(%) unless noted otherwise.
IQR: Interquartile Range. SD: Standard Deviation. cfDNA: cell free fetal DNA.

The prenatally identified 48,XXYY cohort had higher prevalences of several congenital anomalies compared to the general population (Table 2), and the prevalence of cleft lip/palate, hypotonia, and penile chordee/webbing were higher than previously reported for 48,XXYY. This cohort also had higher prevalences of eosinophilic esophagitis (33.3% vs 1.1%; p < 0.001) and allergies (86.7% vs 55.6%; p = 0.025), and lower rates of dental problems (20% vs. 87.6%; p < 0.001) and tremors (20% vs 60.5%; p = 0.014), however, lower rates at the young age of the current cohort will likely increase since these often present later in life. Proportions of cardiac abnormalities and strabismus were similar between those diagnosed prenatally and postnatally.

**Table 2.**
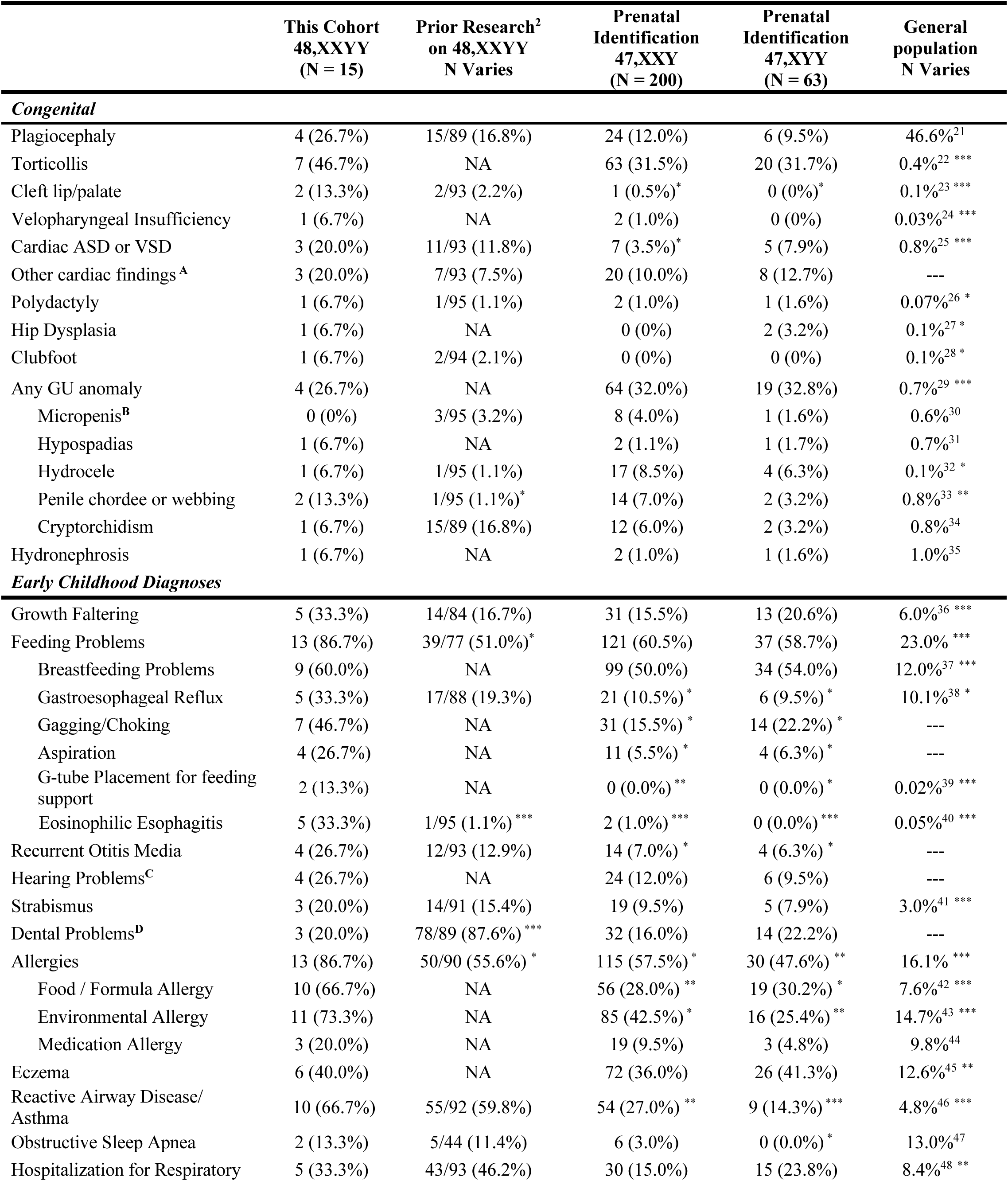
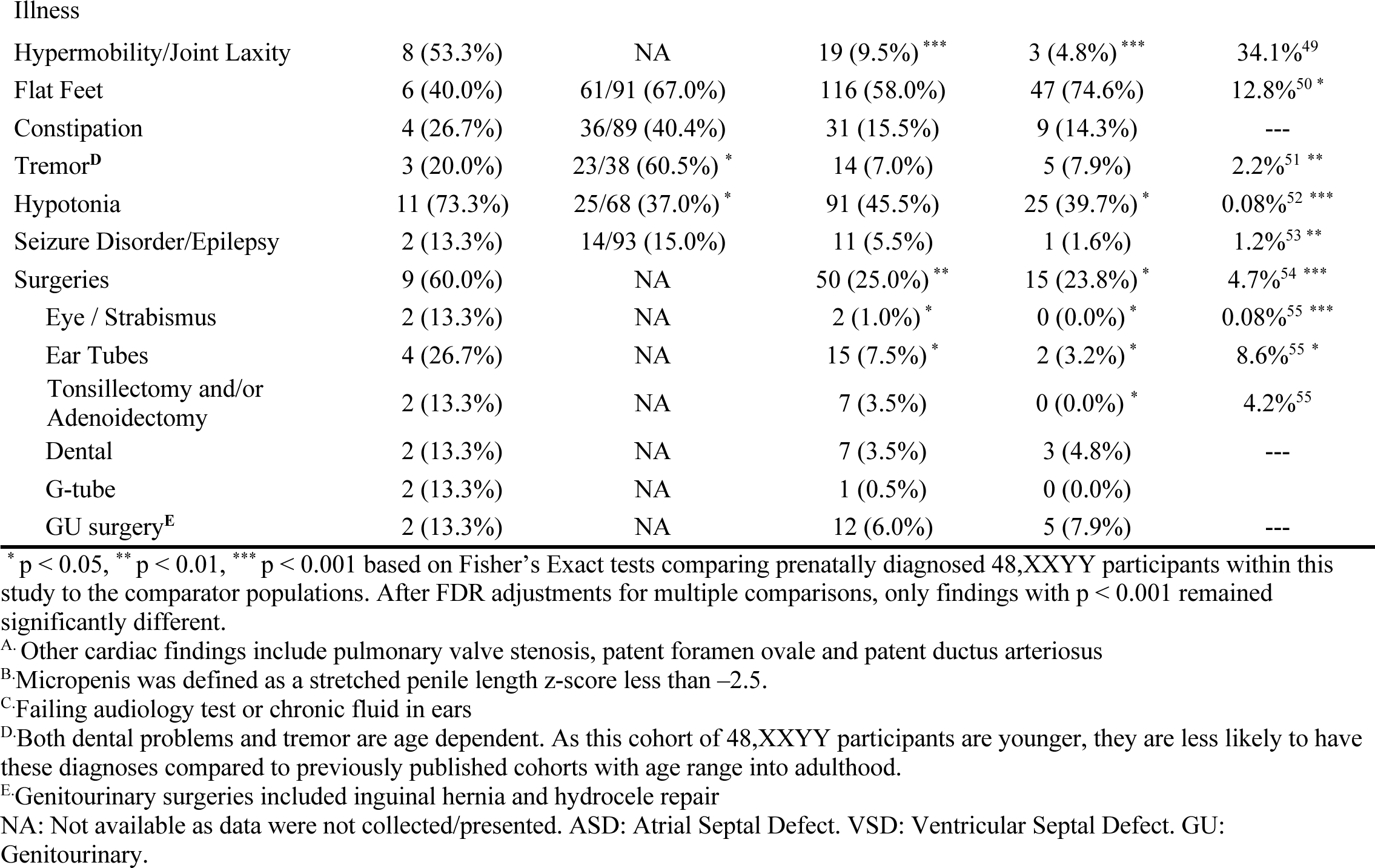
Medical features of prenatally identified 48,XXYY compared to prior research on 48,XXYY, prenatally identified 47,XXY and 47,XYY, and the general population.

Among the 15 participants with 48,XXYY, there was variability in the number of medical conditions between individuals, with some showing few medical diagnoses compared to others with multiple diagnoses (Figure 1). Many conditions also resolved in the first year(s) of life, such as torticollis and gastroesophageal reflux.

**Figure 1.**
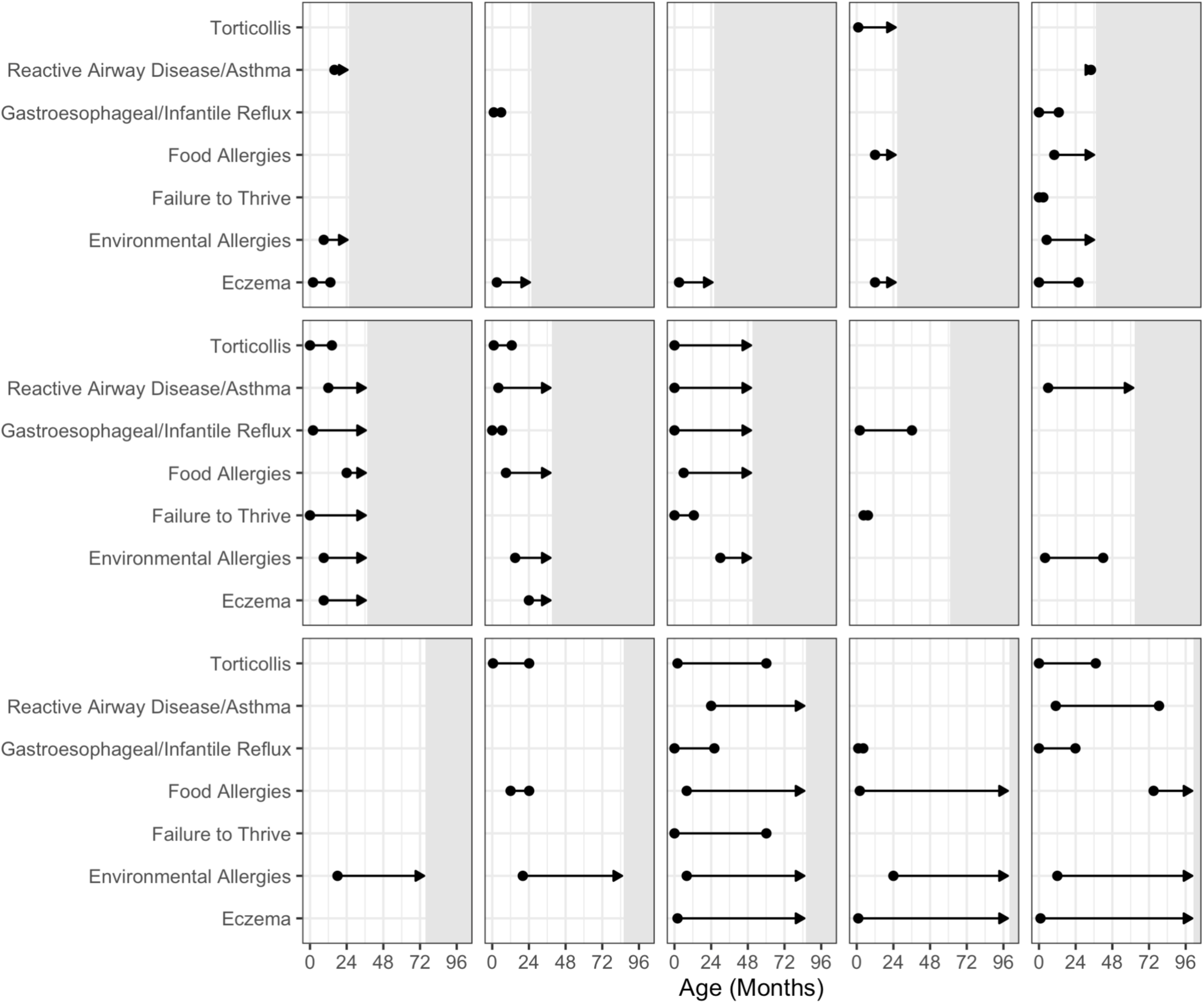
Onset and resolution of select medical features per participant. Points represent the age at the onset and resolution of each condition for each participant, demonstrating variability between participants with some showing few medical problems and others with more medical complexity. Arrowheads indicate that the condition was still present at the most recent follow-up visit, circle indicates it has resolved. Patient ages represent the age, in months, at the most recent follow-up visit. The grey regions represent ages beyond each participant’s final visit, illustrating that participants were too young to contribute data past these points.

Prenatally identified 48,XXYY participants had similar rates of delayed speech and motor milestones compared to previous literature. The proportion of children with 48,XXYY diagnosed with ASD was lower compared to previously reported in XXYY (26.7% vs 52.4%), though not statistically significant. Our cohort also showed higher rates of ASD diagnosis compared to XXY (26.7% vs 5.0%) and XYY (26.7% vs. 4.8%).

The rates of early intervention therapies were very common, and all of the 48,XXYY participants received some type of early intervention therapy (Table 4) started either proactively or due to the presence of developmental delays, with speech, occupational, physical, and feeding therapy the most common therapy types. Compared to both prenatally identified 47,XXY and 47,XYY, a higher proportion of prenatally identified 48,XXYY infants and young children received early intervention therapies.

**Table 3.**
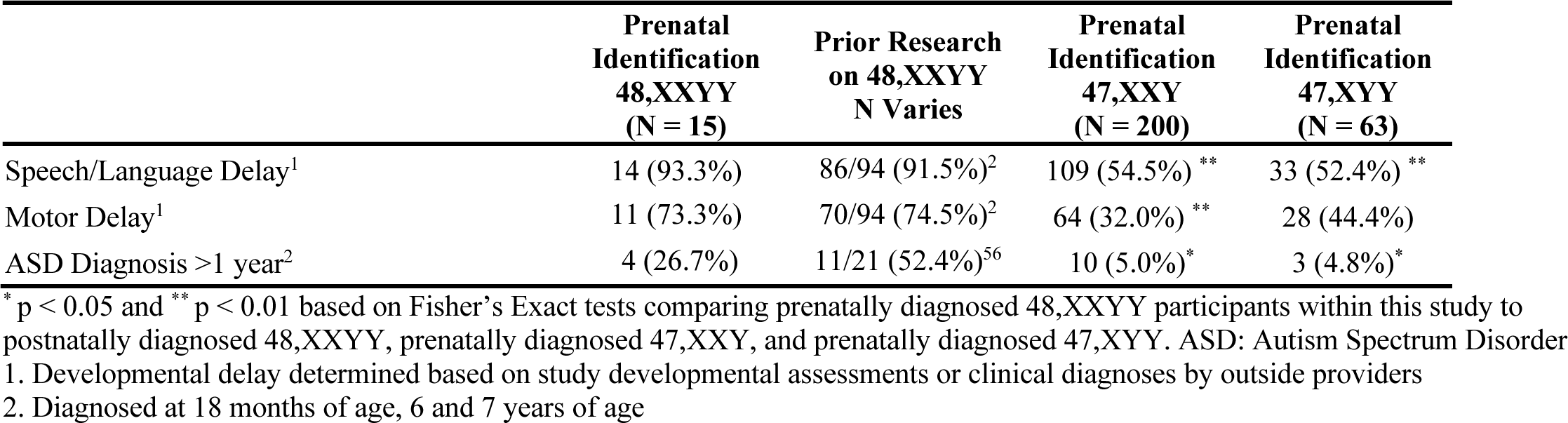
Developmental diagnoses in this prenatally identified 48,XXYY cohort, compared to previous XXYY literature and prenatally identified children with 47,XXY and 47,XYY.

|  | **Prenatal Identification**<br>**48,XXYY**<br>**(N = 15)** | **Prior Research**<br>**on 48,XXYY**<br>**N Varies** | **Prenatal Identification**<br>**47,XXY**<br>**(N = 200)** | **Prenatal Identification**<br>**47,XYY**<br>**(N = 63)** |
| --- | --- | --- | --- | --- |
| Speech/Language Delay<sup>1</sup> | 14 (93.3%) | 86/94 (91.5%)<sup>2</sup> | 109 (54.5%) ** | 33 (52.4%) ** |
| Motor Delay<sup>1</sup> | 11 (73.3%) | 70/94 (74.5%)<sup>2</sup> | 64 (32.0%) ** | 28 (44.4%) |
| ASD Diagnosis >1 year<sup>2</sup> | 4 (26.7%) | 11/21 (52.4%)<sup>56</sup> | 10 (5.0%)* | 3 (4.8%)* |

\* p < 0.05 and \*\* p < 0.01 based on Fisher's Exact tests comparing prenatally diagnosed 48,XXYY participants within this study to postnatally diagnosed 48,XXYY, prenatally diagnosed 47,XXY, and prenatally diagnosed 47,XYY. ASD: Autism Spectrum Disorder
1. Developmental delay determined based on study developmental assessments or clinical diagnoses by outside providers
2. Diagnosed at 18 months of age, 6 and 7 years of age

**Table 4.**
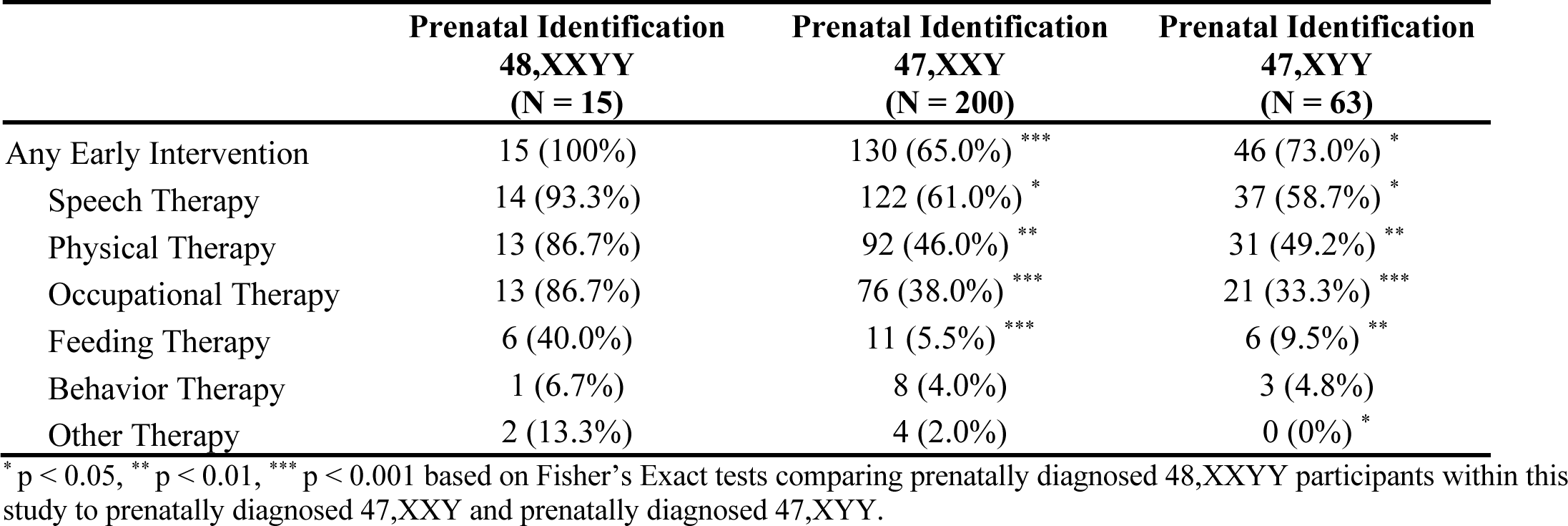
Current or Previous early intervention prevalences.

Developmental milestones were achieved later in participants with 48,XXYY compared to the general population across all domains (Figure 2), however the range at age of achievement is broad and individual data points show many overlapping with the range seen in the general population. Compared to 47,XXY and 47,XYY, the median age of achievement of milestones in 48,XXYY was later for sitting, crawling, cruising, walking, and 2-word phrases (Table 5).

**Figure 2.**
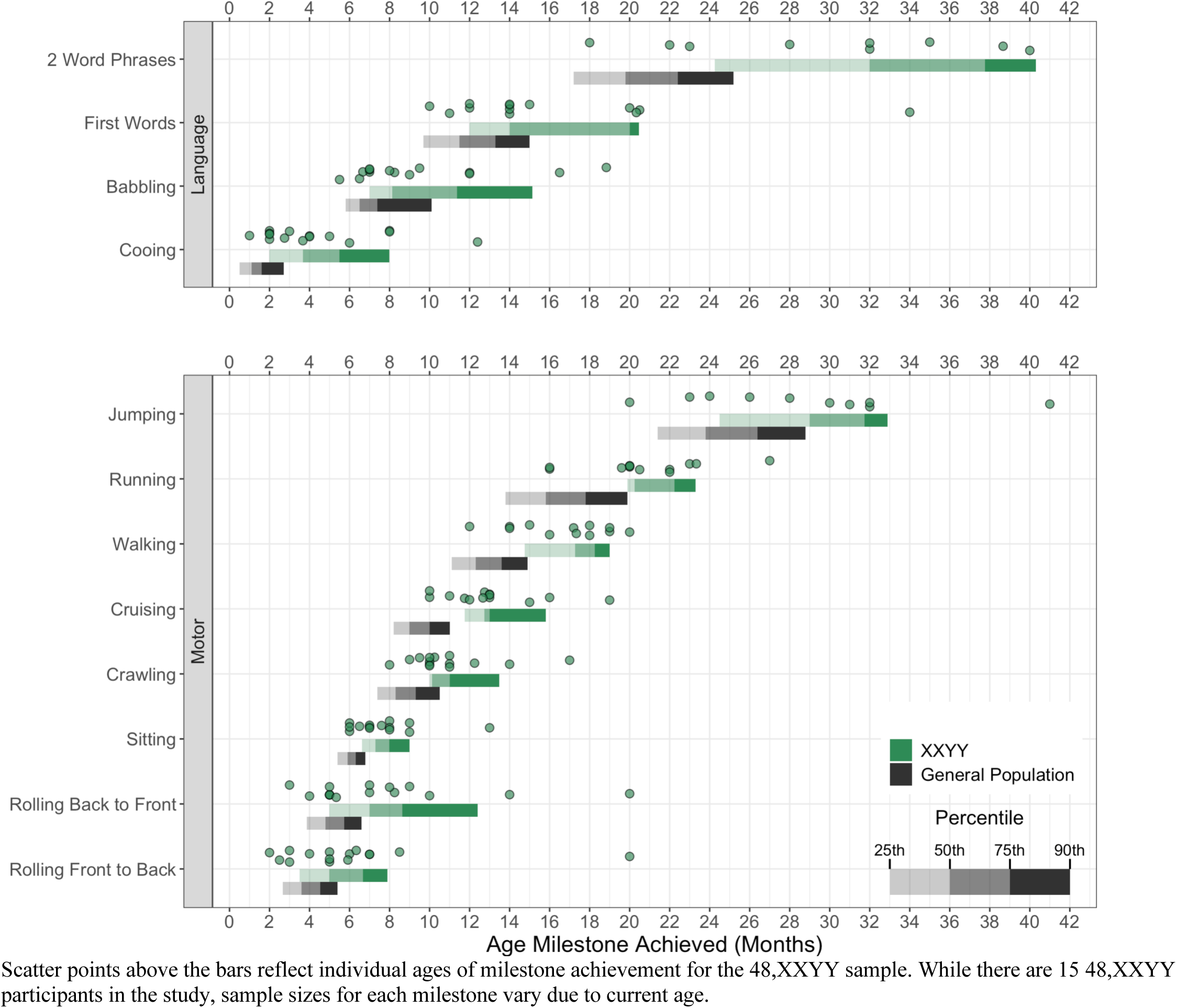
Distribution of 48,XXYY Developmental Milestone Timing. Scatter points above the bars reflect individual ages of milestone achievement for the 48,XXYY sample. While there are 15 48,XXYY participants in the study, sample sizes for each milestone vary due to current age.

**Table 5.**
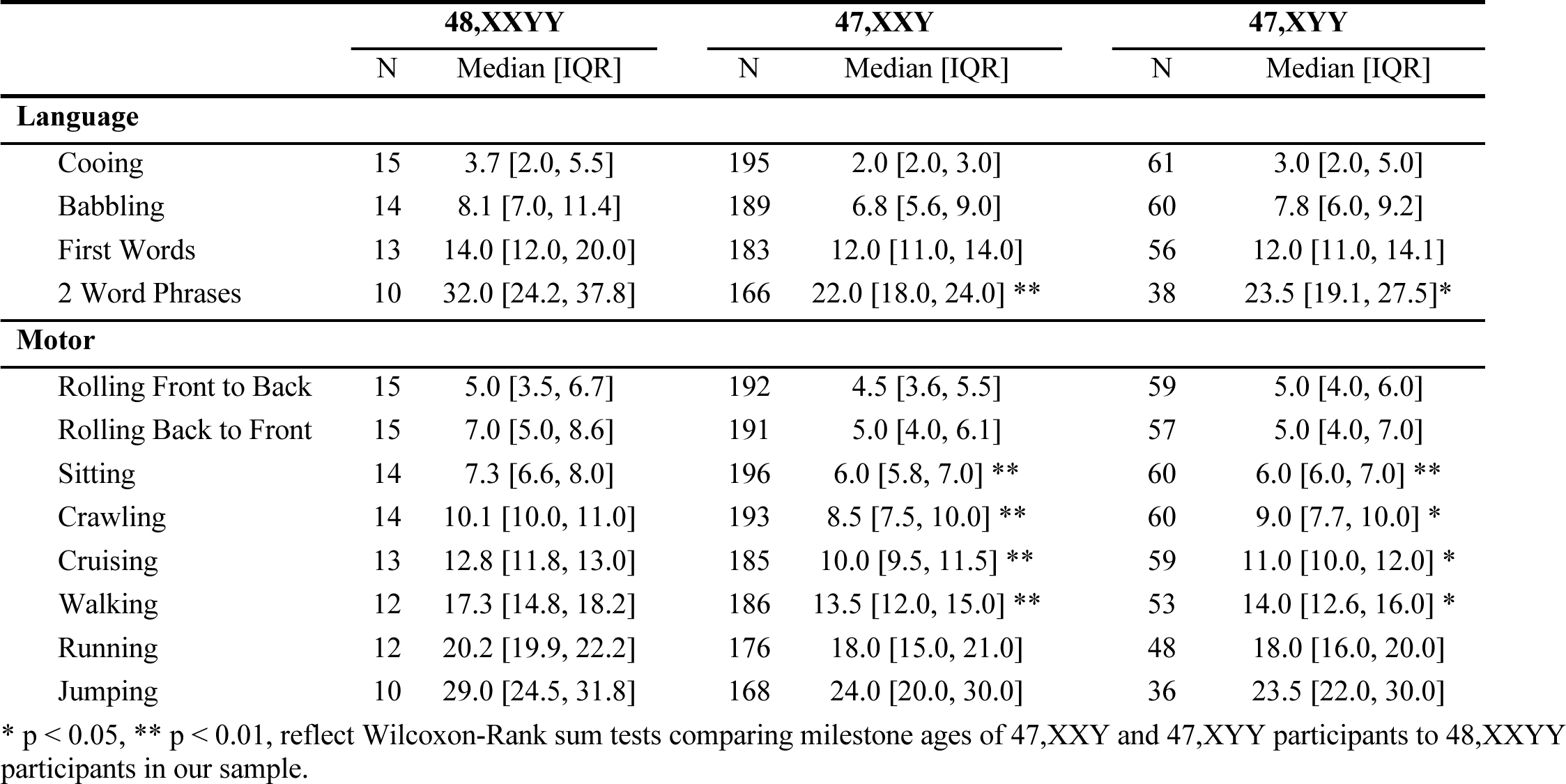
Developmental milestone ages (months) of participants with 48,XXYY compared to those with 47,XXY and 47,XYY.

At 12-months of age, median scores for the 48,XXYY cohort fell within the average to low-average range in all subscales of the Vineland-3 (n=15) and Bayley-III (n=11), with the exception of the Bayley-III gross motor subscale where the median fell in the delayed range (Figure 3). Children with 48,XXYY had lower Vineland-3 adaptive behavior scores compared to 47,XXY and 47,XYY (Figure 3). Longitudinal trajectories of Vineland scores are provided in Supplemental Figure 2 showing trends of decreasing scores compared to general population with advancing age, however sample size decreases at older ages due to some participants not yet reaching those ages. When considering categorical classification of scores, the majority of the 48,XXYY cohort fell in the average range, however was more likely to have impaired adaptive functioning (v-scaled scores ≤9) in expressive communication (20%) and gross motor (33.3%) compared to 2.3% in the general population at 12-months of age. Categorical classification of the 11 48,XXYY participants that completed the 12-month Bayley-III assessment showed that about half fell in the average range and half (45.5%) scored 7 or lower for receptive and/or expressive communication. The gross motor domain was the most significant area of delay, with 8 (73.7%) scoring 7 or lower.

**Figure 3.**
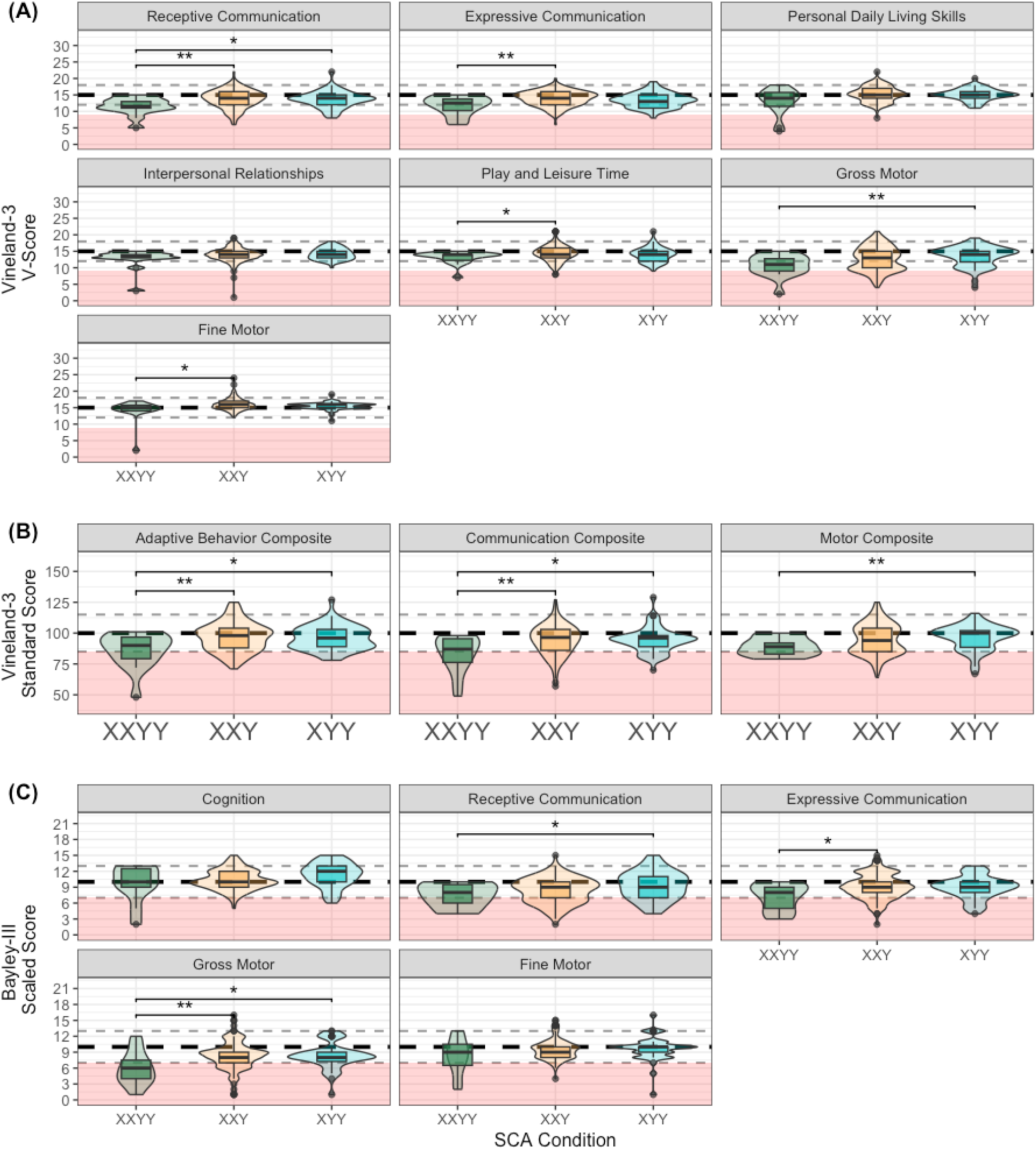
Vineland-3 V-Scores (A), Vineland-3 Composite Standard Scores (B), and Bayley-III Scaled Scores (C) Violin Plot Distributions comparing XXYY to XXY and XYY at 12 Month Visit. * p < 0.05, ** p < 0.01 reflect Wilcoxon-Rank sum tests comparing 48,XXYY Vineland-3 v-scores (A; n=15), Vineland-3 adaptive behavior composite standard scores (B; n=15), and Bayley-III scaled scored (C; n=11) to 47,XXY and 47,XYY at 12 months of age. Boxplots, representing the median and interquartile range, are nested within violin plots. A Vineland-3 v-Score of 15 (A), a Vineland-3 Standard Score of 100 (B), and a Bayley-III scaled score of 10 (C) are considered average and indicated by the black dashed line with the grey lines representing the region within one standard deviation of the mean (3 points for both Vineland-3 v-scores and Bayley-III scaled scores and 15 for the Vineland-3 adaptive behavior composite). The red region reflects below average scores (9 or lower on Vineland) or 7 or lower on Bayley).

## Discussion

This study describes early medical and developmental profiles from the largest prospective cohort of prenatally identified males with 48,XXYY studied to date, addressing a critical gap in a literature that has been based almost entirely on cross-sectional descriptions of clinically ascertained individuals. Our findings support a higher prevalence of many congenital and early childhood health conditions in 48,XXYY compared to both 46,XY males and prenatally identified males with 47,XXY and 47,XYY, as well as substantial variability in developmental milestone attainment. Although many children performed within the average range, the cohort showed later developmental milestone attainment overall and lower developmental scores than prenatally identified cohorts with 47,XXY and 47,XYY. Together, these findings provide important early-life markers that can inform genetic counseling, anticipatory guidance, and clinical care for infants and young children with 48,XXYY.

In line with previous literature on 48,XXYY, our findings confirm increased risk for multisystem medical conditions associated with this condition (Table 2). The prenatal ascertainment and prospective study of this cohort provides new perspective from prior studies, which included largely older and clinically ascertained individuals. This distinction is important because some medical findings, such as dental problems or tremors, typically emerge later in childhood or into adulthood, and may not yet be detectable in our prenatally identified cohort. Conversely, this study highlights several medical features that are prominent in infancy and early childhood that may have been underreported in prior studies of older individuals because of recall limitations or less detailed early-life medical histories. For example, breastfeeding problems, allergies (food / formula and environmental allergies), hypotonia, feeding disorders, torticollis, and eosinophilic esophagitis are all reported at higher rates in this cohort compared to previous studies.

The case series in Figure 1 highlights both the variability and the commonality of medical challenges faced by children with prenatally identified 48,XXYY. Although the specific combination of conditions and severity of conditions differed across individuals, several recurrent patterns emerged. Many children developed respiratory concerns, gastrointestinal difficulties, and atopic conditions. At the same time, there was considerable heterogeneity. Some children experienced feeding problems and growth faltering, while others did not; torticollis was limited to a subset in infancy and resolved with age. Despite this variability, the shared burden of chronic conditions suggests that children with 48,XXYY have overlapping vulnerabilities that may reflect underlying neurodevelopmental, endocrine, and immunologic mechanisms inherent to the aneuploidy.

When considering the neurodevelopmental manifestations of 48,XXYY, our findings generally parallel the pattern observed for medical features: substantial variability and overlap with prenatally identified sex chromosome trisomy cohorts and the general population. Across multiple approaches to characterizing development—including parent-reported adaptive functioning, milestone attainment, continuous Bayley scores, and categorical classification of developmental delay—the overall pattern was consistent. Developmental impact was variable, with some children showing more substantial delays and others demonstrating relatively fewer early developmental concerns.

The milestone data provide practical information for counseling families. The median ages of milestone attainment presented in Table 5, together with Figure 2 displaying quartiles for milestone attainment, help quantify the degree and variability of delay in concrete terms. For example, rather than describing motor delay only in broad terms, these data allow clinicians to explain that children with 48,XXYY may, on average, achieve certain milestones several months later than typically developing children (i.e. independent walking at 17-18 months, about 5 months later than typical children), while also emphasizing the range of outcomes across individuals.

Interpretation of these findings remains limited by the young age of the cohort and the need for continued longitudinal follow-up into later childhood, adolescence, and adulthood. Longer-term data will be essential for defining developmental trajectories, identifying predictors of later outcomes, and determining which early medical, developmental, family, or intervention factors are most informative for prognosis. As additional participants are enrolled and followed over time, estimates of milestone timing and developmental variability will become more precise, strengthening their utility for counseling and clinical planning.

Notably, therapy utilization was higher than rates of documented developmental delay, suggesting that many families and clinicians pursued early intervention proactively rather than waiting for categorical delays to emerge. Understanding how early therapy access and intensity influence long-term outcomes will be important for developing evidence-based recommendations and counseling families about early supports in 48,XXYY.

Comparisons with 47,XXY and 47,XYY cohorts are important because the trisomy conditions themselves are associated with increased medical and developmental risks compared with the general population.^57,58^ In the present study, however, children with 48,XXYY showed an even greater burden of several medical and developmental concerns, suggesting a potential dosage effect of additional sex chromosome material. This stepwise increase in clinical involvement from sex chromosome trisomy to tetrasomy may provide a useful framework for future gene expression studies, as the molecular pathways disrupted in 47,XXY and 47,XYY may be even more dysregulated in 48,XXYY. Such studies could help clarify which sex chromosome dosage-sensitive genes or downstream pathways contribute to shared and syndrome-specific features across sex chromosome aneuploidies.

Although prenatal ascertainment reduces referral bias and should theoretically capture a wider range of outcomes, the medical and developmental findings in this cohort were largely consistent with those reported in clinically ascertained 48,XXYY cohorts. This contrasts somewhat with sex chromosome trisomy research (XXY, XYY, XXX), where prenatal versus postnatal ascertainment often has a substantial impact on observed phenotype severity.^59,60,61^ In 48,XXYY, the presence of both an additional X and Y chromosome results in more pronounced dosage-related effects, such that core features are observed even in cohorts identified prenatally.

These findings have important implications for both prenatal and postnatal genetic counseling. Almost all participants in this study had prenatal cfDNA results reported as positive for a sex chromosome trisomy (XXY or XYY), and experienced counseling based only on information about sex chromosome trisomy, which underrepresents the medical and developmental risks in 48,XXYY. Prevalence of the more complex medical conditions in 48,XXYY and the higher likelihood of neurodevelopmental disorders necessitates different genetic counseling for XXYY syndrome. These results reinforce previous recommendations for counselors to include possibility of a more complex SCA condition when counseling for positive prenatal SCA cfDNA results,^14,62^ and offering prenatal genetic confirmatory testing (chorionic villi sampling or amniocentesis) if definitive diagnosis is important to the family. If XXYY syndrome is confirmed in the prenatal period or in infancy, genetic counselors can play an important role in helping families anticipate early referrals, advocate for developmental services, connect with community advocacy organizations, and establish appropriate medical monitoring after birth (Table 3). The variability in outcomes emphasizes the importance of personalized care plans for 48,XXYY patients and ongoing developmental assessments for early identification of emerging concerns to optimize outcomes.

These results should also guide medical care for infants with XXYY. Primary care providers should be aware of the common medical findings in XXYY syndrome so that appropriate screenings and medical evaluations can be implemented. In particular, careful evaluation for congenital anomalies, including a postnatal echocardiogram, is pertinent for early identification and appropriate management. Previous publications from the eXtraordinarY Babies Study include recommendations for primary care providers for management of sex chromosome trisomy conditions, which can also be useful for XXYY.^63^

**Table 5.**
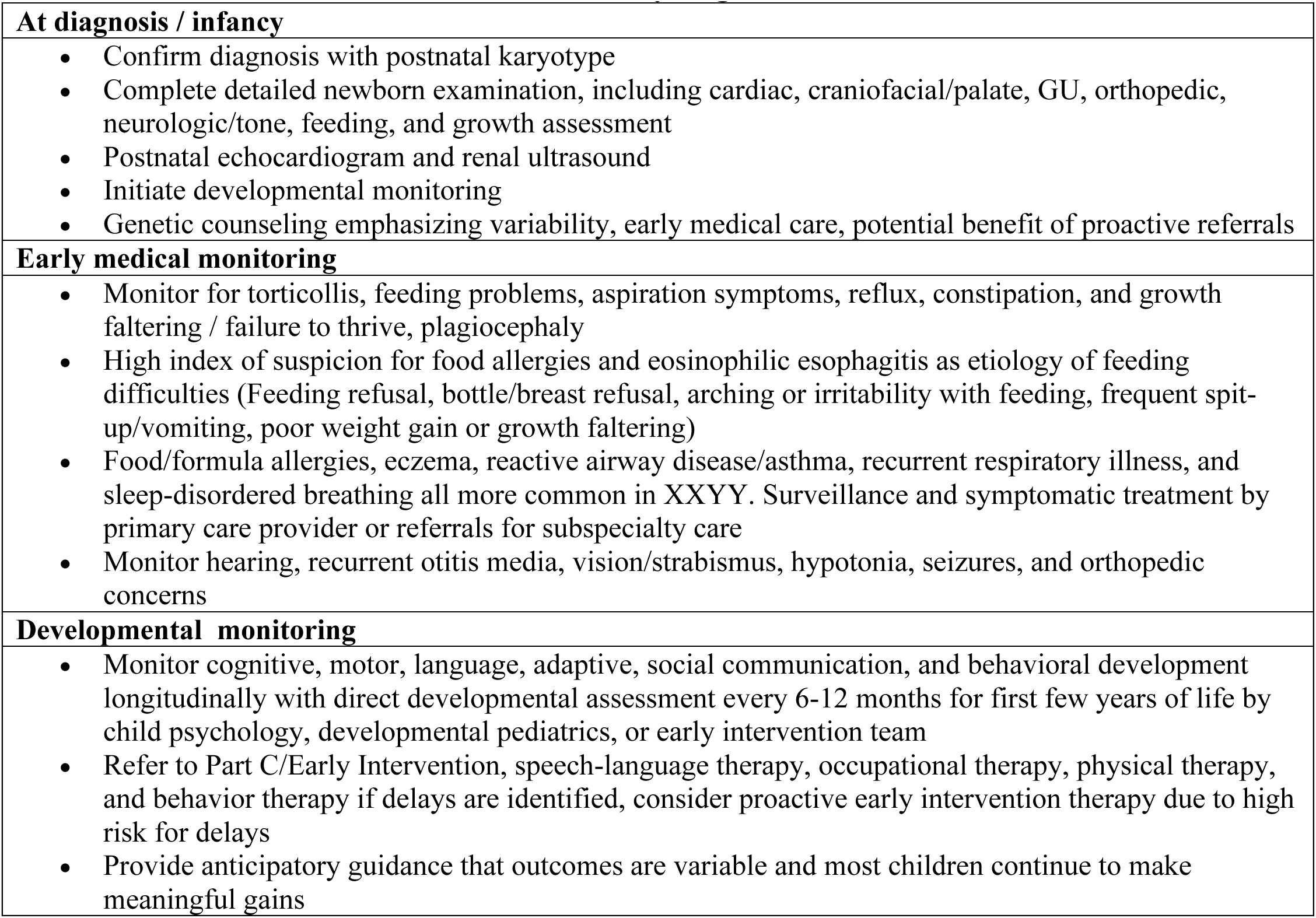
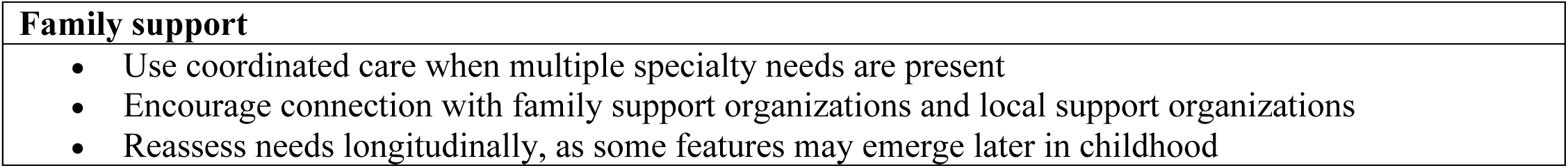
Clinical care considerations for infants and young children with 48,XXYY.

|  |
| --- |
| **At diagnosis / infancy** |
| • Confirm diagnosis with postnatal karyotype<br>• Complete detailed newborn examination, including cardiac, craniofacial/palate, GU, orthopedic, neurologic/tone, feeding, and growth assessment<br>• Postnatal echocardiogram and renal ultrasound<br>• Initiate developmental monitoring<br>• Genetic counseling emphasizing variability, early medical care, potential benefit of proactive referrals |
| **Early medical monitoring** |
| • Monitor for torticollis, feeding problems, aspiration symptoms, reflux, constipation, and growth faltering / failure to thrive, plagiocephaly<br>• High index of suspicion for food allergies and eosinophilic esophagitis as etiology of feeding difficulties (Feeding refusal, bottle/breast refusal, arching or irritability with feeding, frequent spit-up/vomiting, poor weight gain or growth faltering)<br>• Food/formula allergies, eczema, reactive airway disease/asthma, recurrent respiratory illness, and sleep-disordered breathing all more common in XXYY. Surveillance and symptomatic treatment by primary care provider or referrals for subspecialty care<br>• Monitor hearing, recurrent otitis media, vision/strabismus, hypotonia, seizures, and orthopedic concerns |
| **Developmental monitoring** |
| • Monitor cognitive, motor, language, adaptive, social communication, and behavioral development longitudinally with direct developmental assessment every 6-12 months for first few years of life by child psychology, developmental pediatrics, or early intervention team<br>• Refer to Part C/Early Intervention, speech-language therapy, occupational therapy, physical therapy, and behavior therapy if delays are identified, consider proactive early intervention therapy due to high risk for delays<br>• Provide anticipatory guidance that outcomes are variable and most children continue to make meaningful gains |

Several limitations should be considered. Although this is the largest prospective cohort of prenatally identified boys with 48,XXYY reported to date, the sample size remains small secondary to the rarity of the condition. This limits our power to detect statistical differences where differences may exist, and may not fully capture the variability of medical and developmental outcomes that would be seen in a larger and more diverse sample. Additionally, given our young cohort, our findings do not capture medical or neurodevelopmental problems that appear later in life, and ongoing longitudinal follow-up will be essential to characterize the full developmental and medical trajectory of 48,XXYY across the lifespan. Also, our study cohort is biased towards families with higher levels of education and socioeconomic status who have access to more specialized medical care and who may be more likely to access early intervention services.

## Conclusions

In conclusion, this study provides the first detailed prospective characterization of early medical and developmental outcomes in prenatally identified boys with 48,XXYY. Although prenatal ascertainment may reduce referral bias and capture a broader phenotypic spectrum, the findings were broadly consistent with prior reports of clinically ascertained individuals, suggesting that core features of 48,XXYY are evident early in life and remain clinically important even when the diagnosis is made prenatally. Compared with prenatally identified 47,XXY and 47,XYY cohorts, boys with 48,XXYY were more likely to show a more complex early phenotype, including higher rates of several congenital, feeding, respiratory, allergic/immunologic, gastrointestinal, neurologic, and neurodevelopmental concerns, agreeing with the previously coined phrase that 48,XXYY is “not just a variant of Klinefelter syndrome.”^4^. At the same time, substantial variability was observed, reinforcing the need for individualized care. These findings support diagnosis-specific anticipatory guidance, proactive medical and developmental surveillance beginning in infancy, and coordinated multidisciplinary care (Table 5). Continued longitudinal follow-up of these children will be essential in refining our understanding of the developmental pathways associated with 48,XXYY. Continued longitudinal follow-up of these children will be important to define later-emerging features, identify predictors of outcomes, and refine evidence-based care recommendations across the lifespan. and improving outcomes through targeted interventions.

## Data Availability

This study is registered on ClinicalTrials.gov NCT03396562 https://clinicaltrials.gov/study/NCT03396562. Data sharing is available through requests from NICHD DASH (https://dash.nichd.nih.gov/) and includes deidentified individual participant data, study protocols, DASH data codebook, and the informed consent form. The data will be made available to researchers who provide methodologically sound proposals approved by the NICHD DASH Data or Biospecimen Access Committee.

## Abbreviations

SCA: Sex Chromosome Aneuploidy
ASD: autism spectrum disorder
ADHD: attention-deficit/hyperactivity disorder
cfDNA: Cell free fetal DNA

---

**Supplemental Figure 1.**
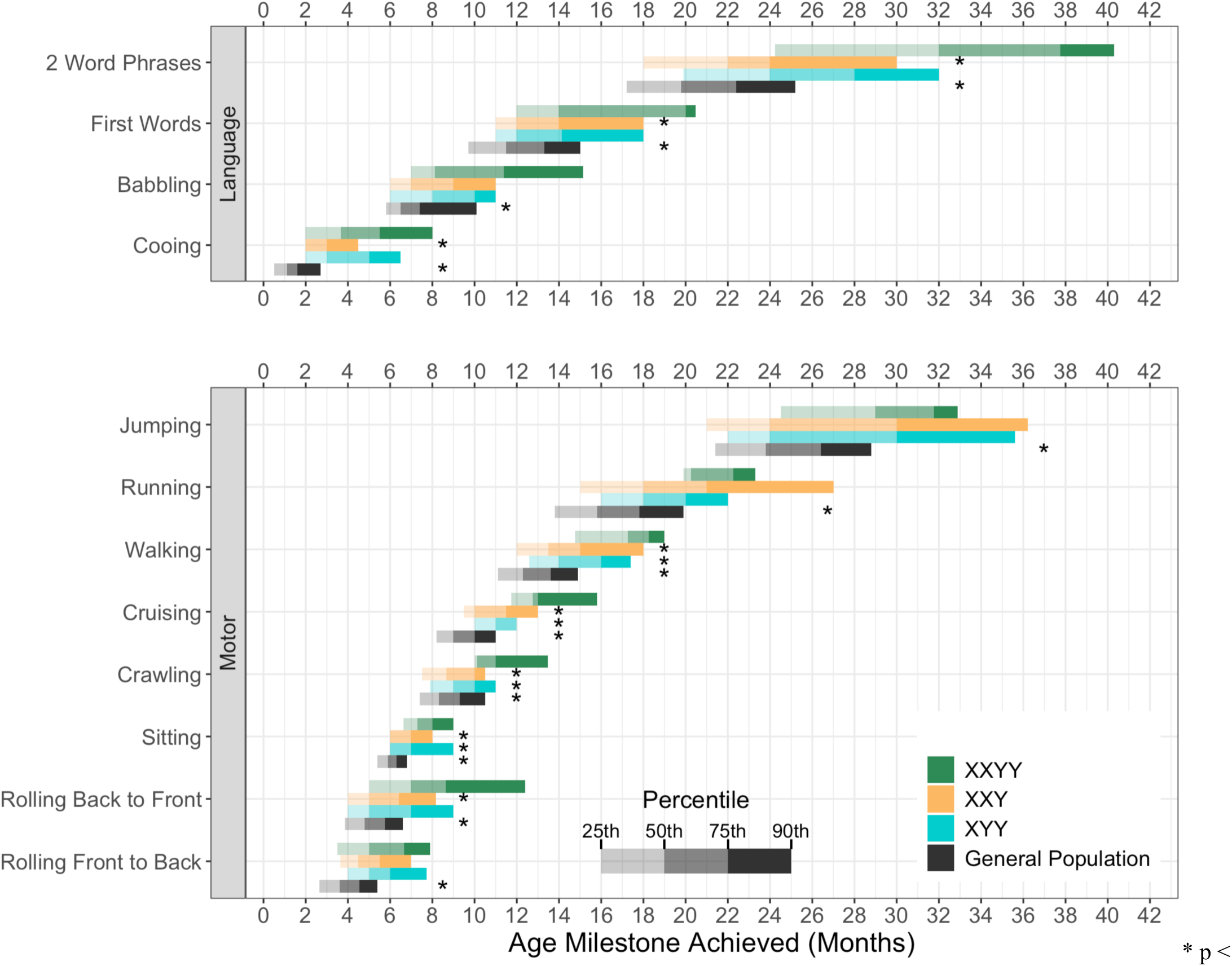
Distribution of Sex Chromosome Aneuploidy Developmental Milestone Timing Compared to the General Population. * p < 0.05 reflect Wilcoxon-Rank sum tests comparing milestone ages of 48,XXYY to 47,XXY, 47,XYY, simulated data based on general population percentiles, taken from the Denver II scales, the World Health Organization (WHO) Motor Development Study, and the Primitive Reflex Profile (PRP), under the assumption of a non-normal distribution.^15-17^

**Supplemental Figure 2.**
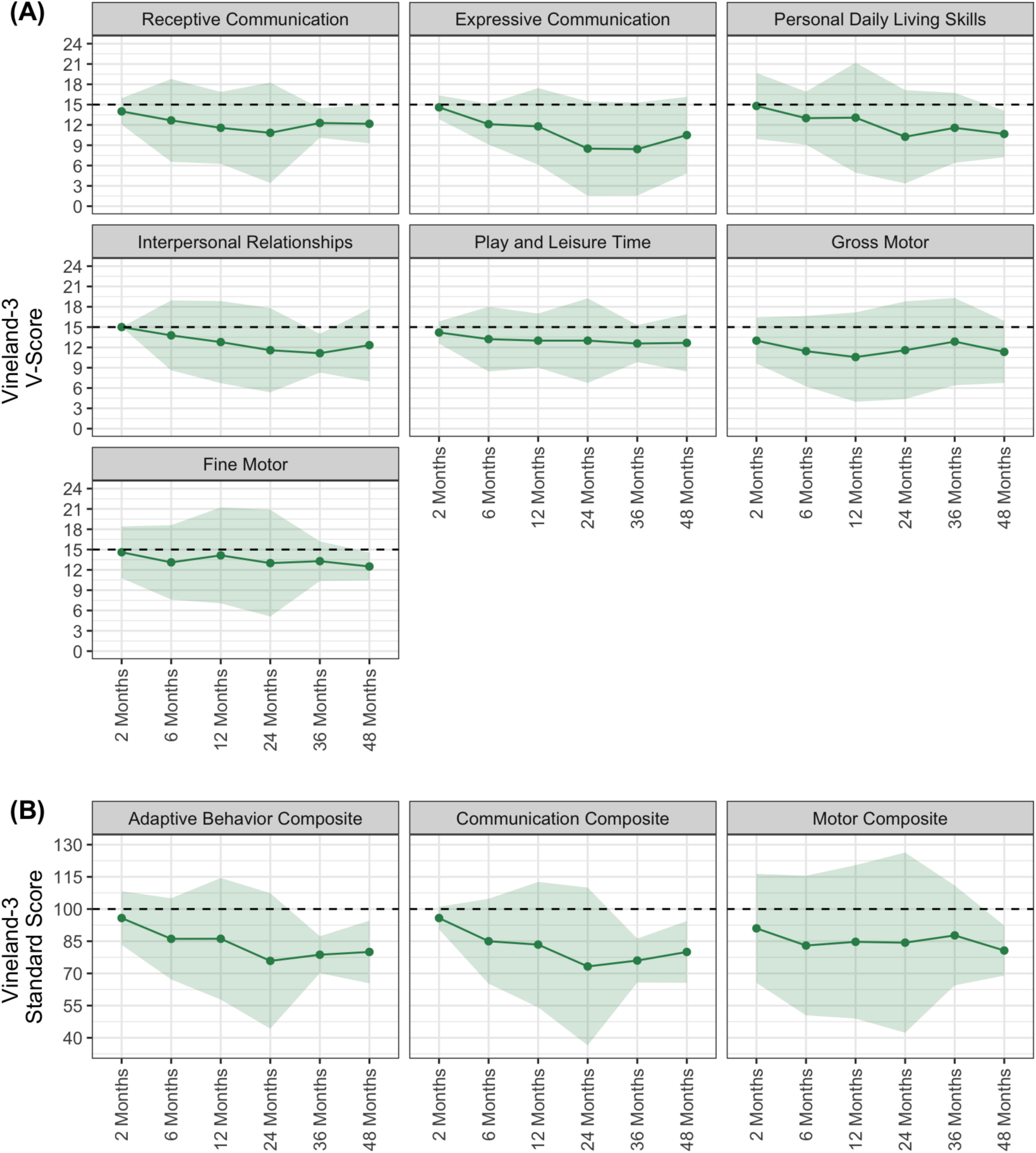
Vineland-3 Longitudinal 48,XXYY Scores. Trends in Vineland-3 adaptive scores from 2 to 48 months of age, indicating achievement of adaptive skills not keeping pace with typical expectations. Data points from all available data included, although n’s decrease with age due to some participants not yet reaching those ages (12m n=15; 24m n=14; 36m n=7; 48m n=7; 48m n=4). Points and ribbons reflect mean and 95% confidence intervals for scores, respectively.

